# Association between Prolonged Intermittent Renal Replacement Therapy and All-Cause Mortality in COVID-19 Patients Undergoing Invasive Mechanical Ventilation: a Retrospective Cohort Study

**DOI:** 10.1101/2020.03.16.20036780

**Authors:** Yi Yang, Jia Shi, Shuwang Ge, Shuiming Guo, Xue Xing, Yanan Wang, Anying Cheng, Qingquan Liu, Junhua Li, Yong Ning, Fan He, Gang Xu

**Author notes:** These authors contributed equally to this work (Yi Yang, Jia Shi, Shuwang Ge). Correspondence Author: Fan He and Gang Xu, Department of Nephrology, Tongji Hospital Affiliated with Tongji Medical College, Huazhong University of Science and Technology, 1095 Jie Fang Avenue, Wuhan, Hubei 430030, China, and.

## Abstract

**Background:** For the coronavirus disease 2019 (COVID-19), critically ill patients had a high mortality rate. We aimed to assess the association between prolonged intermittent renal replacement therapy (PIRRT) and mortality in patients with COVID-19 undergoing invasive mechanical ventilation.

**Methods:** In this retrospective cohort study, we included all patients with COVID-19 undergoing invasive mechanical ventilation from February 12^nd^ to March 2^nd^, 2020. All patients were followed until death or March 28^th^, and all survivors were followed for at least 30 days.

**Results:** For 36 hospitalized COVID-19 patients with invasive mechanical ventilation, the mean age was 69.4 (± 10.8) years, and 30 patients (83.3%) were men. Twenty-two (61.1%) patients received PIRRT (PIRRT group) and 14 cases (38.9%) were managed with conventional strategy (non-PIRRT group). There were no differences in age, sex, comorbidities, complications, treatments and most of the laboratory findings. During median follow-up period of 9.5 (interquartile range 4.3-33.5) days, 13 of 22 (59.1%) patients in the PIRRT group and 11 of 14 (78.6%) patients in the non-PIRRT group died. Kaplan-Meier analysis demonstrated prolonged survival in patients in the PIRRT group compared with that in the non-PIRRT group (P = 0.042). The association between PIRRT and a reduced risk of mortality remained significant in three different models, with adjusted hazard ratios varying from 0.332 to 0.398. Higher levels of IL-2 receptor, TNF-α, procalcitonin, prothrombin time, and NT-proBNP were significantly associated with an increased risk of mortality in patients with PIRRT.

**Conclusion:** PIRRT may be beneficial for the treatment of COVID-19 patients with invasive mechanical ventilation. Further prospective multicenter studies with larger sample sizes are required.

## Introduction

An outbreak of severe acute respiratory syndrome coronavirus 2 (SARS-CoV-2) infection, officially named Coronavirus Disease 2019 (COVID-19) occurred (1). In Wuhan, the fatality rate of COVID-19 was 5.1% (2538/50006). Of note, critically ill patients with COVID-19 have a high mortality rate. In a study of 52 critically ill patients in Wuhan, 32 (61.5%) patients had died after 28 days, and the mortality rate was 81.1% (30/37) in patients requiring mechanical ventilation (2). Accumulated evidence has strongly demonstrated that systemic inflammatory response, acute kidney injury (AKI) and fluid overload (FO) were associated with high mortality in severe sepsis (3-5). In critically ill patients with COVID-19, an overwhelming inflammatory response involving C-reactive protein (CRP), interleukin (IL)-6, IL-8, IL-10, tumor necrosis factor *α* (TNF-α) was observed (6-9), which is similar to that observed in patients suffering from SARS-CoV (10) and Middle East respiratory syndrome (MERS)-CoV (11).

Renal replacement therapy (RRT) is a great help in treating critically ill patients not only to control electrolyte and acid-base imbalances but also to remove inflammatory mediators and improve oxygenation during fluid overload (12-14). RRT has been applied to critically ill patients, including patients with SARS-CoV, MERS-CoV and other viral infectious diseases such as Ebola virus disease (13, 15). However, the benefits of RRT are still no consistent conclusion in critically ill patients (16). RRT significantly reduced the level of IL-6 and decreased the hospital mortality rate in pediatric severe sepsis, especially in patients with acute respiratory distress syndrome (ARDS) (17). In addition, a meta-analysis revealed that patients who received RRT had significantly lower mortality compared to conventional therapy (18). Hoverer, RRT was associated with increased mortality in patients with MERS-CoV (15). The relationship between RRT and patient outcome varied in patients with different disease and was affected by the modalities, use of anticoagulation, vascular access management, and the timing of the initiation and intensity of RRT (16,19,20).

Prolonged intermittent renal replacement therapy (PIRRT), as a cost-effective alternative, has been used in the intensive care unit (ICU) (21, 22). To date, no specific treatment has been confirmed to be effective for COVID-19, and supportive treatment remains essential. In this retrospective cohort study, we aimed to explore the association between PIRRT and all-cause mortality in patients with COVID-19 undergoing invasive mechanical ventilation.

## Materials and Methods

### Study Design and Participants

In this retrospective cohort study, we included all patients with COVID-19 undergoing invasive mechanical ventilation at the Optical Valley Branch of Tongji Hospital, Wuhan, from February 12^nd^ to March 2^nd^. We divided the study participants into two groups according to the use of PIRRT treatment (PIRRT group and non-PIRRT group).

### Inclusion and Exclusion Criteria

All included patients met the criteria for the diagnosis of COVID-19 according to the New Coronavirus Pneumonia Prevention and Control Program (5^th^ edition, in Chinese) published by the National Health Commission of China (23). Invasive mechanical ventilation was defined as mechanical ventilation through an endotracheal tube or tracheostomy. There were no exclusion criteria.

### Procedures

The baseline data at the beginning of invasive mechanical ventilation for each patient were collected and recorded at the start of invasive mechanical ventilation, including age, sex, comorbidities, complications, laboratory data, and treatments. All information was obtained and managed through established data collection forms. Two researchers independently reviewed and collected the data. AKI was defined as a 50% increase in serum creatinine within 7 days or a 0.3 mg/dL increase within 48 hours according to the Kidney Disease: Improving Global Outcomes (KDIGO) criteria (24).

### PIRRT Procedures

We performed PIRRT using commercially available pump-driven machines (PrismaFlex, Gambro, Sweden; or multiFiltrate, Fresenius, Germany) and filters (M150, Gambro; Oxiris, Baxter; or AVIOOOs, Fresenius). For the patients with AKI, hémofiltration plus hemodialysis was performed. The blood flow rate was set at 2.5-4 mL/kg/min, and the clearance rate was set at 35-70 mL/kg/hr. The filter circuit was prewashed with saline containing 5,000-6,250 IU/L heparin. Vascular access was obtained with 13.5 F central venous catheters (Covidien, MA, USA) in the femoral vein. 9.1% (2/22) of patients received continuous hemodialysis. And 90.9% (20/22) of patients received intermittent hemodialysis, 8 hours a day, once a day or every other day. PIRRT modalities were venovenous hemodiafiltration in 22.7% (5/22) of patients and venovenous hemofiltration in 77.3% (17/22) of patients.

The indications for PIRRT was: 1) Nonobstructive oliguria (urine output < 200 mL/12h) or anuria, or sepsis complicated by AKI; 2) Hyperkalemia (K+ > 6.5 mmol/L); 3) Acidemia (PH < 7.1); 4) Clinically significant organ edema (especially pulmonary edema); 5) Uremic complications (pericarditis/encephalopathy/neuropathy/myopathy); 6) Azotaemia (urea > 30 mmol/L); 7) (optional) increased inflammatory cytokines (anyone of IL-1β, IL-2 receptor, IL-6, IL-8, or TNF-α ≥ 5 times of upper limit of normal range).

### Outcomes

We followed up all patients through electronic hospital medical records. The primary outcome was death. All patients were followed until death or March 28^th^, and all survivors were followed for at least 30 days. There was no loss to follow-up for patients.

### Statistical Analyses

Numerical data are presented as the means and standard deviation (SD) or medians [interquartile range (IQR)] and were analyzed using Student’s t-test or the Mann-Whitney U test depending on the data distribution. Categorical variables were displayed as frequencies and percentages and were analyzed using Fisher’s exact test. Paired t test or Wilcoxon matched-pairs signed rank test was used to evaluate differences of variables between before and after PIRRT. The Kaplan-Meier method was used to estimate survival, and the log-rank test was used to evaluate differences between the two groups. Univariate and multivariate Cox proportional hazards regression analyses were performed for all-cause mortality. SPSS 23.0 (IBM Corporation, Armonk, NY) statistical software was used for statistical analysis. GraphPad Prism 6 (GraphPad Software, USA) was used for statistical analysis and visualization. A P-value of 0.05 or less was considered significant.

## Results

### Description of the Cohort

In total, 36 COVID-19 patients subjected to invasive mechanical ventilation were enrolled in the study. Table 1 shows the baseline characteristics of the cohort patients. There were 30 men (83.3%) and 6 women (16.7%) ranging in age from 44 to 86 years. 30 patients (83.3%) have at least one comorbidity, and the common comorbidity factors in COVID-19 patients with invasive mechanical ventilation were hypertension (n = 14, 38.9%) and cardiac disease (n = 12, 33.3%).

**Table 1.**
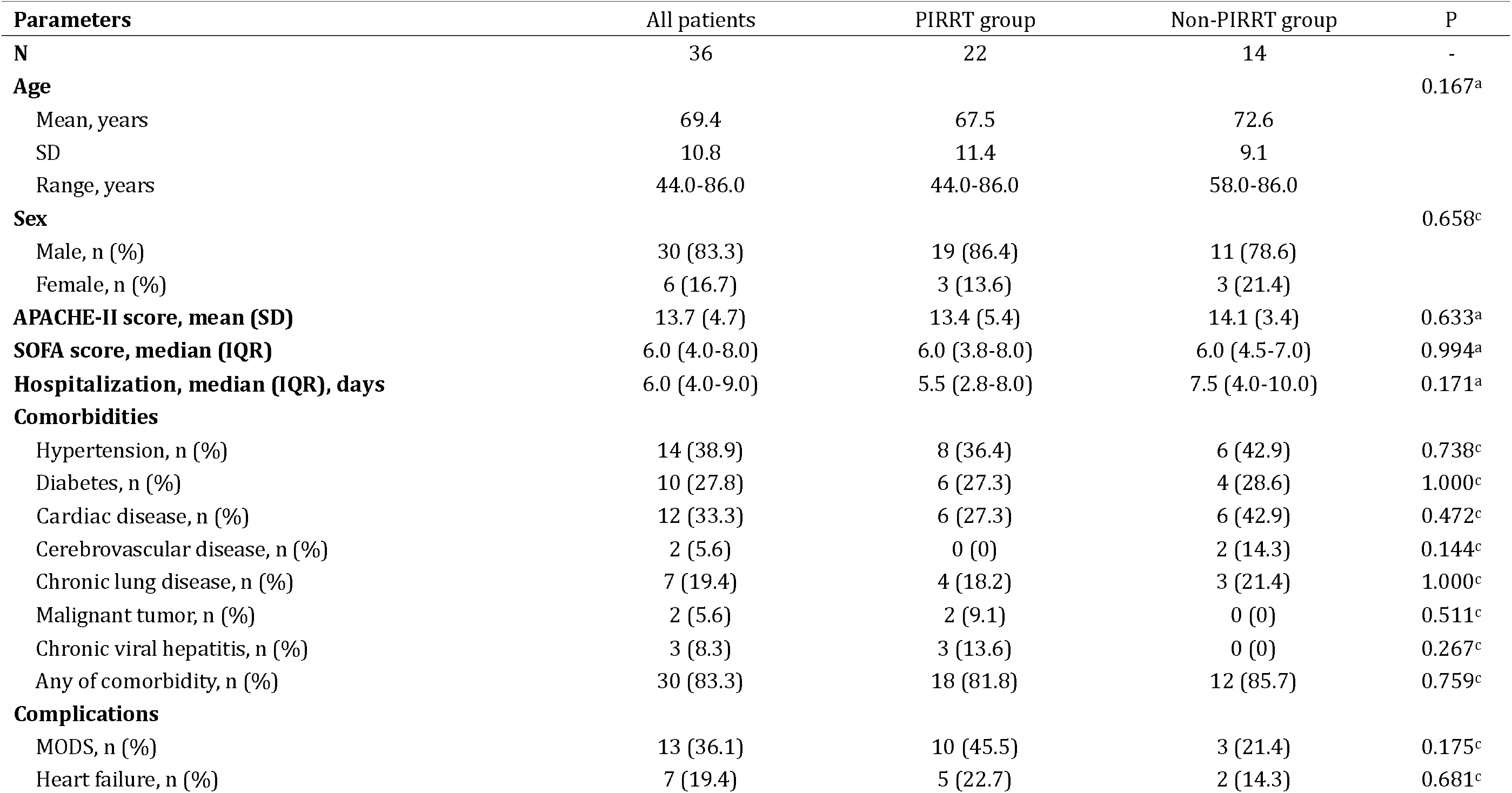
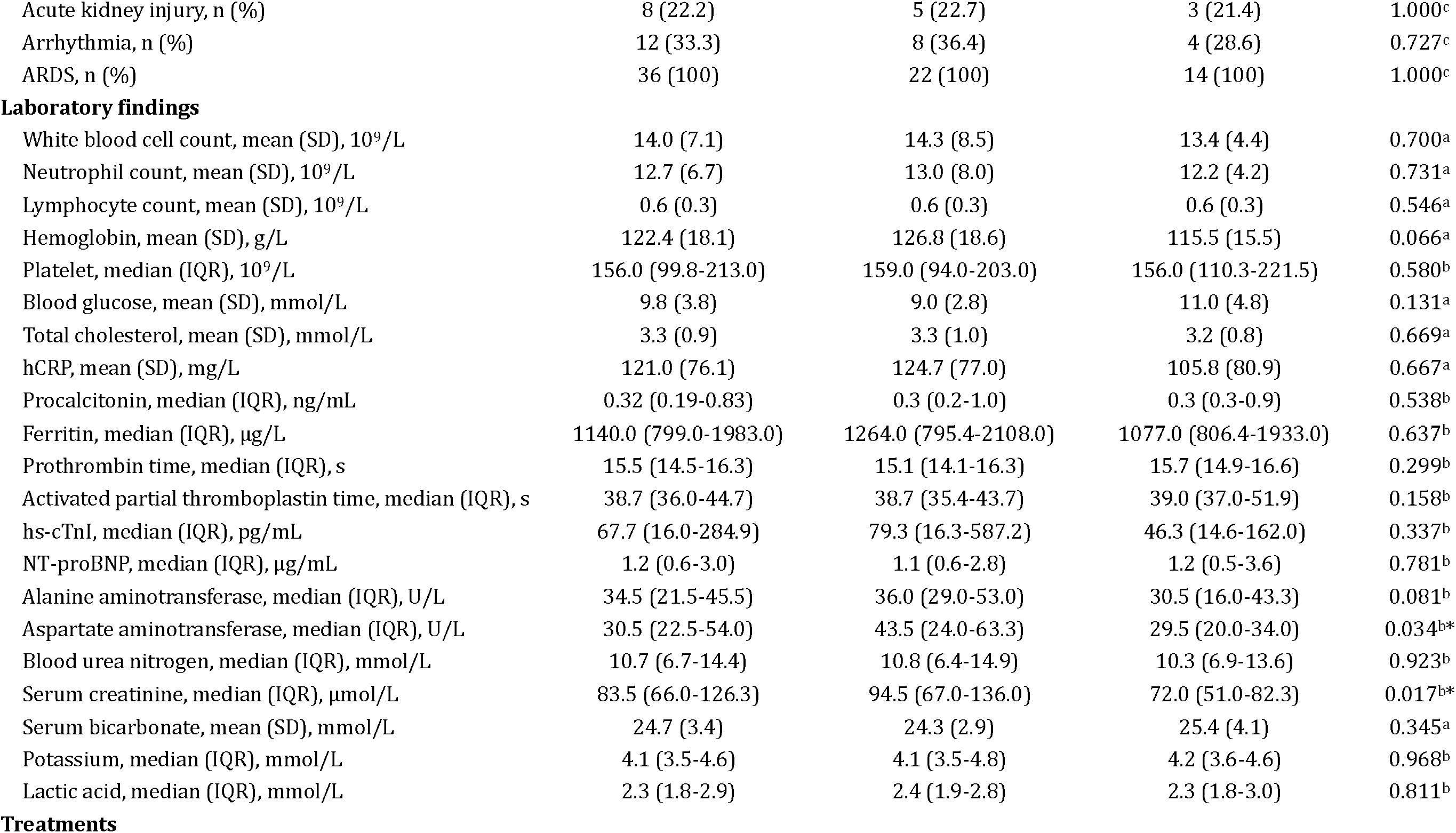
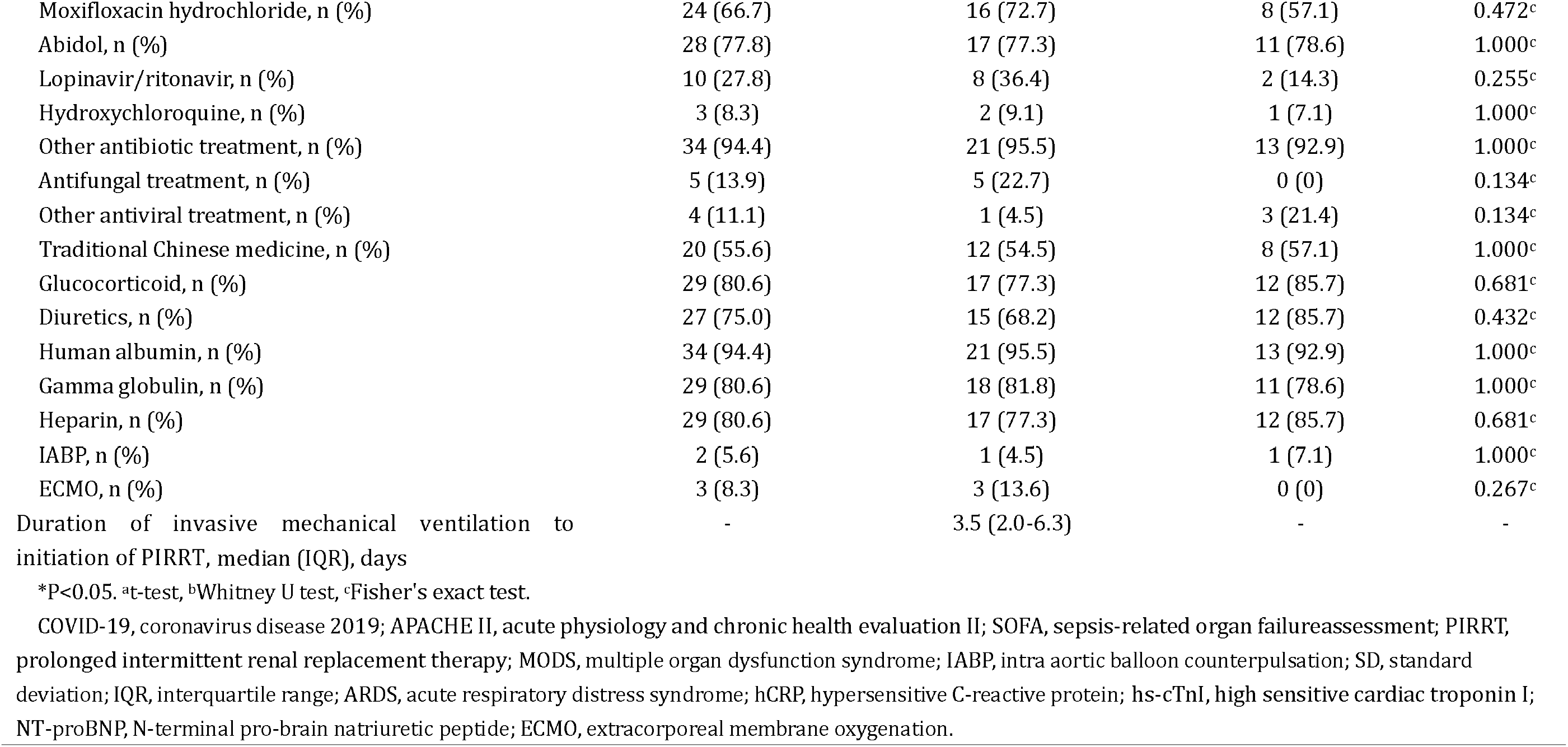
Comparison of baseline demographics and clinical characteristics of COVID-19 patients undergoing invasive mechanical ventilation between patients with and without PIRRT treatment in the cohort

| Parameters | All patients | PIRRT group | Non-PIRRT group | P |
| --- | --- | --- | --- | --- |
| **N** | 36 | 22 | 14 | - |
| **Age** |  |  |  | 0.167<sup>a</sup> |
| Mean, years | 69.4 | 67.5 | 72.6 |  |
| SD | 10.8 | 11.4 | 9.1 |  |
| Range, years | 44.0-86.0 | 44.0-86.0 | 58.0-86.0 |  |
| **Sex** |  |  |  | 0.658<sup>c</sup> |
| Male, n (%) | 30 (83.3) | 19 (86.4) | 11 (78.6) |  |
| Female, n (%) | 6 (16.7) | 3 (13.6) | 3 (21.4) |  |
| **APACHE-II score, mean (SD)** | 13.7 (4.7) | 13.4 (5.4) | 14.1 (3.4) | 0.633<sup>a</sup> |
| **SOFA score, median (IQR)** | 6.0 (4.0-8.0) | 6.0 (3.8-8.0) | 6.0 (4.5-7.0) | 0.994<sup>a</sup> |
| **Hospitalization, median (IQR), days** | 6.0 (4.0-9.0) | 5.5 (2.8-8.0) | 7.5 (4.0-10.0) | 0.171<sup>a</sup> |
| **Comorbidities** |  |  |  |  |
| Hypertension, n (%) | 14 (38.9) | 8 (36.4) | 6 (42.9) | 0.738<sup>c</sup> |
| Diabetes, n (%) | 10 (27.8) | 6 (27.3) | 4 (28.6) | 1.000<sup>c</sup> |
| Cardiac disease, n (%) | 12 (33.3) | 6 (27.3) | 6 (42.9) | 0.472<sup>c</sup> |
| Cerebrovascular disease, n (%) | 2 (5.6) | 0 (0) | 2 (14.3) | 0.144<sup>c</sup> |
| Chronic lung disease, n (%) | 7 (19.4) | 4 (18.2) | 3 (21.4) | 1.000<sup>c</sup> |
| Malignant tumor, n (%) | 2 (5.6) | 2 (9.1) | 0 (0) | 0.511<sup>c</sup> |
| Chronic viral hepatitis, n (%) | 3 (8.3) | 3 (13.6) | 0 (0) | 0.267<sup>c</sup> |
| Any of comorbidity, n (%) | 30 (83.3) | 18 (81.8) | 12 (85.7) | 0.759<sup>c</sup> |
| **Complications** |  |  |  |  |
| MODS, n (%) | 13 (36.1) | 10 (45.5) | 3 (21.4) | 0.175<sup>c</sup> |
| Heart failure, n (%) | 7 (19.4) | 5 (22.7) | 2 (14.3) | 0.681<sup>c</sup> |
| Acute kidney injury, n (%) | 8 (22.2) | 5 (22.7) | 3 (21.4) | 1.000<sup>c</sup> |
| Arrhythmia, n (%) | 12 (33.3) | 8 (36.4) | 4 (28.6) | 0.727<sup>c</sup> |
| ARDS, n (%) | 36 (100) | 22 (100) | 14 (100) | 1.000<sup>c</sup> |
| **Laboratory findings** |  |  |  |  |
| White blood cell count, mean (SD), 10<sup>9</sup>/L | 14.0 (7.1) | 14.3 (8.5) | 13.4 (4.4) | 0.700<sup>a</sup> |
| Neutrophil count, mean (SD), 10<sup>9</sup>/L | 12.7 (6.7) | 13.0 (8.0) | 12.2 (4.2) | 0.731<sup>a</sup> |
| Lymphocyte count, mean (SD), 10<sup>9</sup>/L | 0.6 (0.3) | 0.6 (0.3) | 0.6 (0.3) | 0.546<sup>a</sup> |
| Hemoglobin, mean (SD), g/L | 122.4 (18.1) | 126.8 (18.6) | 115.5 (15.5) | 0.066<sup>a</sup> |
| Platelet, median (IQR), 10<sup>9</sup>/L | 156.0 (99.8-213.0) | 159.0 (94.0-203.0) | 156.0 (110.3-221.5) | 0.580<sup>b</sup> |
| Blood glucose, mean (SD), mmol/L | 9.8 (3.8) | 9.0 (2.8) | 11.0 (4.8) | 0.131<sup>a</sup> |
| Total cholesterol, mean (SD), mmol/L | 3.3 (0.9) | 3.3 (1.0) | 3.2 (0.8) | 0.669<sup>a</sup> |
| hCRP, mean (SD), mg/L | 121.0 (76.1) | 124.7 (77.0) | 105.8 (80.9) | 0.667<sup>a</sup> |
| Procalcitonin, median (IQR), ng/mL | 0.32 (0.19-0.83) | 0.3 (0.2-1.0) | 0.3 (0.3-0.9) | 0.538<sup>b</sup> |
| Ferritin, median (IQR), µg/L | 1140.0 (799.0-1983.0) | 1264.0 (795.4-2108.0) | 1077.0 (806.4-1933.0) | 0.637<sup>b</sup> |
| Prothrombin time, median (IQR), s | 15.5 (14.5-16.3) | 15.1 (14.1-16.3) | 15.7 (14.9-16.6) | 0.299<sup>b</sup> |
| Activated partial thromboplastin time, median (IQR), s | 38.7 (36.0-44.7) | 38.7 (35.4-43.7) | 39.0 (37.0-51.9) | 0.158<sup>b</sup> |
| hs-cTnI, median (IQR), pg/mL | 67.7 (16.0-284.9) | 79.3 (16.3-587.2) | 46.3 (14.6-162.0) | 0.337<sup>b</sup> |
| NT-proBNP, median (IQR), µg/mL | 1.2 (0.6-3.0) | 1.1 (0.6-2.8) | 1.2 (0.5-3.6) | 0.781<sup>b</sup> |
| Alanine aminotransferase, median (IQR), U/L | 34.5 (21.5-45.5) | 36.0 (29.0-53.0) | 30.5 (16.0-43.3) | 0.081<sup>b</sup> |
| Aspartate aminotransferase, median (IQR), U/L | 30.5 (22.5-54.0) | 43.5 (24.0-63.3) | 29.5 (20.0-34.0) | 0.034<sup>b*</sup> |
| Blood urea nitrogen, median (IQR), mmol/L | 10.7 (6.7-14.4) | 10.8 (6.4-14.9) | 10.3 (6.9-13.6) | 0.923<sup>b</sup> |
| Serum creatinine, median (IQR), µmol/L | 83.5 (66.0-126.3) | 94.5 (67.0-136.0) | 72.0 (51.0-82.3) | 0.017<sup>b*</sup> |
| Serum bicarbonate, mean (SD), mmol/L | 24.7 (3.4) | 24.3 (2.9) | 25.4 (4.1) | 0.345<sup>a</sup> |
| Potassium, median (IQR), mmol/L | 4.1 (3.5-4.6) | 4.1 (3.5-4.8) | 4.2 (3.6-4.6) | 0.968<sup>b</sup> |
| Lactic acid, median (IQR), mmol/L | 2.3 (1.8-2.9) | 2.4 (1.9-2.8) | 2.3 (1.8-3.0) | 0.811<sup>b</sup> |
| **Treatments** |  |  |  |  |
| Moxifloxacin hydrochloride, n (%) | 24 (66.7) | 16 (72.7) | 8 (57.1) | 0.472<sup>c</sup> |
| Abidol, n (%) | 28 (77.8) | 17 (77.3) | 11 (78.6) | 1.000<sup>c</sup> |
| Lopinavir/ritonavir, n (%) | 10 (27.8) | 8 (36.4) | 2 (14.3) | 0.255<sup>c</sup> |
| Hydroxychloroquine, n (%) | 3 (8.3) | 2 (9.1) | 1 (7.1) | 1.000<sup>c</sup> |
| Other antibiotic treatment, n (%) | 34 (94.4) | 21 (95.5) | 13 (92.9) | 1.000<sup>c</sup> |
| Antifungal treatment, n (%) | 5 (13.9) | 5 (22.7) | 0 (0) | 0.134<sup>c</sup> |
| Other antiviral treatment, n (%) | 4 (11.1) | 1 (4.5) | 3 (21.4) | 0.134<sup>c</sup> |
| Traditional Chinese medicine, n (%) | 20 (55.6) | 12 (54.5) | 8 (57.1) | 1.000<sup>c</sup> |
| Glucocorticoid, n (%) | 29 (80.6) | 17 (77.3) | 12 (85.7) | 0.681<sup>c</sup> |
| Diuretics, n (%) | 27 (75.0) | 15 (68.2) | 12 (85.7) | 0.432<sup>c</sup> |
| Human albumin, n (%) | 34 (94.4) | 21 (95.5) | 13 (92.9) | 1.000<sup>c</sup> |
| Gamma globulin, n (%) | 29 (80.6) | 18 (81.8) | 11 (78.6) | 1.000<sup>c</sup> |
| Heparin, n (%) | 29 (80.6) | 17 (77.3) | 12 (85.7) | 0.681<sup>c</sup> |
| IABP, n (%) | 2 (5.6) | 1 (4.5) | 1 (7.1) | 1.000<sup>c</sup> |
| ECMO, n (%) | 3 (8.3) | 3 (13.6) | 0 (0) | 0.267<sup>c</sup> |
| Duration of invasive mechanical ventilation to initiation of PIRRT, median (IQR), days | - | 3.5 (2.0-6.3) | - | - |

\*P<0.05. <sup>a</sup>t-test, <sup>b</sup>Whitney U test, <sup>c</sup>Fisher's exact test.
COVID-19, coronavirus disease 2019; APACHE II, acute physiology and chronic health evaluation II; SOFA, sepsis-related organ failure assessment; PIRRT, prolonged intermittent renal replacement therapy; MODS, multiple organ dysfunction syndrome; IABP, intra aortic balloon counterpulsation; SD, standard deviation; IQR, interquartile range; ARDS, acute respiratory distress syndrome; hCRP, hypersensitive C-reactive protein; hs-cTnI, high sensitive cardiac troponin I; NT-proBNP, N-terminal pro-brain natriuretic peptide; ECMO, extracorporeal membrane oxygenation.

We divided the study participants into two groups according to PIRRT treatment. Twenty-two patients received PIRRT (PIRRT group) while 14 patients did not (non-PIRRT group). There was no difference between the two groups in baseline characteristics including age, sex, acute physiology and chronic health evaluation (APACHE) II scores, sepsis-related organ failure assessment (SOFA) scores, comorbidities, complications, treatments or most laboratory findings, except for patients who received PIRRT with higher levels of aspartate aminotransferase (P = 0.034) and serum creatinine (P = 0.017).

The indications for PIRRT (n = 22) were as follows: (1) AKI at stage 3 (serum creatinine increase ≥ 3 times baseline with 7 days) with/without hyperkalemia or pulmonary edema: n = 4; (2) hyperkalemia: n = 1; (3) acidemia: n = 1; (4) pulmonary edema: n = 1; (5) (optional) increased inflammatory cytokines (anyone of IL-1β, IL-2 receptor, IL-6, IL-8, or TNF-α ≥ 5 times of upper limit of normal range): n = 15. There was no definite indication of PIRRT for patients in non-PIRRT group. Our evaluation of dialysis indications was consistent in all patients, except for inflammatory cytokines.

Furthermore, in the PIRRT group, IL-6 showed a significant difference before versus after PIRRT (before vs. after PIRRT: median 221.35, IQR 111.23-427.40, vs. median 48.53, IQR 12.93-119.23, pg/mL, P = 0.001).

#### Association between PIRRT and All-Cause Mortality in COVID-19 Patients Undergoing Invasive Mechanical Ventilation

All survivors were followed for at least 30 days. During the median follow-up period of 9.5 (IQR 4.3-33.5) days, 13 of 22 (59.1%) patients in the PIRRT group and 11 of 14 (78.6%) patients in the non-PIRRT group died. Kaplan-Meier analysis indicated that the patients in the PIRRT group had prolonged survival compared to those in the non-PIRRT group (P = 0.04) (shown in Fig. 1).

**Fig. 1.**
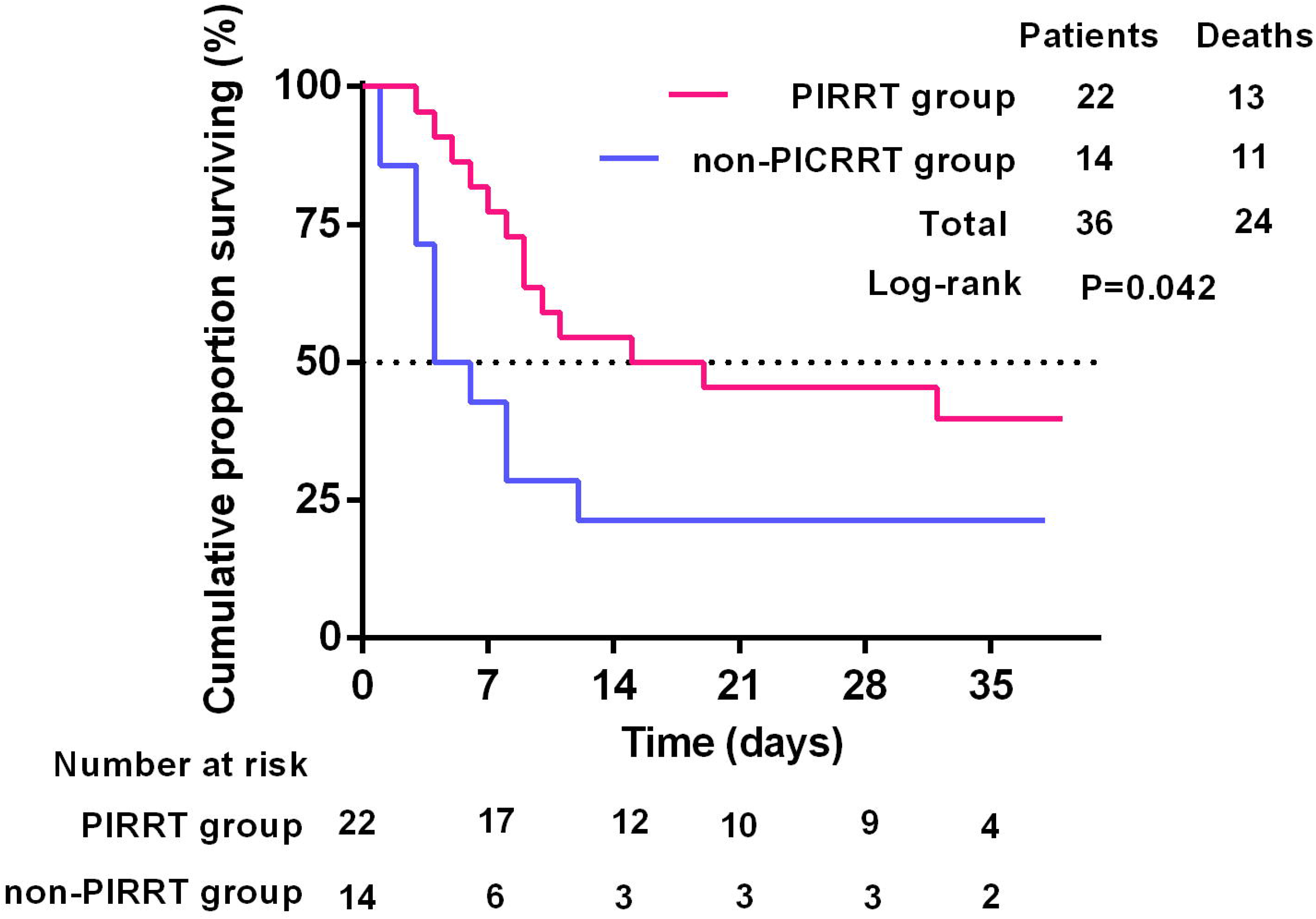
Kaplan-Meier curve of overall patient survival according to with or without PIRRT treatment. Patient survival was significantly better for PIRRT group than for non-PIRRT group (log-rank test, P=0.042).

In the Cox regression analysis, three different models were used to analyze the adjusted hazard ratio for PIRRT treatment. Consistently, the association between PIRRT treatment and a reduced risk of mortality remained significant and the adjusted hazard ratio (aHRs) of PIRRT treatment fluctuated between 0.332 and 0.398 (Table 2).

**Table 2.** Models of multivariate Cox proportional hazard regression analysis for PIRRT treatment (reference group: non-PIRRT treatment) for all-cause mortality of all COVID-19 patients undergoing invasive mechanical ventilation in the cohort

| PIRRT versus non-PIRRT | Adjusted Hazard Ratio (95% CI) | P |
| --- | --- | --- |
| **Model A** | 0.350 (0.147, 0.830) | 0.017 |
| **Model B** | 0.398 (0.171, 0.924) | 0.032 |
| **Model C** | 0.332 (0.119, 0.925) | 0.035 |
| Model A: Adjusted for APACHE II scores, SOFA scores, and any of comorbidity; |  |  |
| Model B: Adjusted for Acute kidney injury, APACHE II scores, Sex; |  |  |
| Model C: Adjusted for IL-6, APACHE II scores, IL-2 receptor. |  |  |

#### Risk Factors Associated with All-Cause Mortality for COVID-19 Patients Undergoing Invasive Mechanical Ventilation with PIRRT Treatment

We further conducted univariate Cox proportional hazard regression analyses of all-cause mortality of COVID-19 patients undergoing invasive mechanical ventilation with PIRRT. We found that higher levels of IL-2 receptor, TNF-α, procalcitonin, prothrombin time, and NT-proBNP were significantly associated with an increased risk of all-cause mortality (Table 3).

**Table 3.** Univariate Cox proportional hazard regression analysis for all-cause mortality of COVID-19 patients undergoing invasive mechanical ventilation with PIRRT treatment in the cohort

| Parameters | Hazard Ratio (95% CI) | P |
| --- | --- | --- |
| Age (per year) | 1.048 (0.998, 1.100) | 0.059 |
| Sex (male vs. female) | 0.959 (0.209, 4.397) | 0.957 |
| APACHE II score (per 1 score) | 1.038 (0.942, 1.143) | 0.453 |
| SOFA score (per 1 score) | 1.093 (0.948, 1.261) | 0.221 |
| Any of comorbidity (with vs. without) | 1.720 (0.379, 7.816) | 0.483 |
| Acute kidney injury (with vs. without) | 2.219 (0.674, 7.313) | 0.190 |
| White blood cell count (per $10^9/L$) | 0.980 (0.918, 1.047) | 0.553 |
| IL-2 receptor (per U/mL) | 1.002 (1.001, 1.003) | 0.004* |
| TNF-α (per pg/mL) | 1.046 (1.002, 1.092) | 0.041* |
| Procalcitonin (per ng/L) | 2.306 (1.098, 4.842) | 0.027* |
| Prothrombin time (per s) | 1.808 (1.229, 2.659) | 0.003* |
| D-dimer (per $\mu g/mL$ [FEU]) | 1.034 (0.964, 1.109) | 0.346 |
| hs-cTnI (per pg/mL) | 1.000 (1.000, 1.000) | 0.257 |
| NT-proBNP (per $\mu g/mL$) | 1.181 (1.056, 1.320) | 0.003* |
| Alanine aminotransferase (per U/L) | 0.998 (0.992, 1.004) | 0.479 |
| Aspartate aminotransferase (per U/L) | 0.998 (0.994, 1.003) | 0.442 |
| Plasma albumin (per g/L) | 1.010 (0.856, 1.191) | 0.910 |
| Blood urea nitrogen (per mmol/L) | 1.021 (0.950, 1.099) | 0.569 |
| Serum creatinine (per $\mu mol/L$) | 1.000 (0.987, 1.012) | 0.963 |

COVID-19, coronavirus disease 2019; PIRRT, prolonged intermittent renal replacement therapy; APACHE II, acute physiology and chronic health evaluation II; SOFA, sepsis-related organ failure assessment; IL, interleukin; TNF, tumor necrosis factor; FEU, fibrinogen equivalent units; hs-cTnI, high sensitive cardiac troponin I; NT-proBNP, N-terminal pro-brain natriuretic peptide.

## Discussion

To the best of our knowledge, this study is the first cohort study to estimate the association between PIRRT treatment and the mortality of COVID-19 patients subjected to invasive mechanical ventilation. We included 36 COVID-19 patients subjected to invasive mechanical ventilation, of whom 22 patients received PIRRT. During the follow-up, 59.1% of patients in the PIRRT group, and 78.6% of patients in the non-PIRRT group died. PIRRT was independently associated with prolonged survival and a lower risk of mortality in COVID-19 patients requiring invasive mechanical ventilation.

Excessive inflammation characterized by the uncontrolled release of pro-inflammatory cytokines into circulation is the main cause of death from sepsis (25, 26), and infection with influenza virus (27), Ebola virus (28), MERS-CoV (29), and SARS-CoV (30). In our study, we found that the cytokine storm might play a crucial role in critical COVID-19 patients. The mean/median levels of inflammatory markers, including IL-1β, IL-2 receptor, IL-6, IL-8, and IL-10, white blood cell count, neutrophil count, hCRP, procalcitonin, and ferritin were higher than normal. And in our study, univariate Cox proportional hazard regression analysis showed increased IL-6 [HR=1.004, 95% CI (1.001-1.007)] and TNF-α [HR=1.040, 95% CI (1.007-1.073)] were both associated with increased risk of mortality of all patients with invasive mechanical ventilation. In patients undergoing PIRRT, IL-6 showed significant difference before versus after PIRRT: before vs. after PIRRT: 221.35 (IQR 111.23-427.40) vs. 48.53 (IQR 12.93-119.23), P=0.001.

Cytokine storms may be caused by the following aspects. First, SARS-CoV-2 infects patients by binding human angiotensin (Ang)-converting enzyme 2 (ACE2) (31, 32), which is widely expressed in multiple organs throughout the body (33). SARS-CoV-2 might lead to multisystem inflammation through the ACE/Ang II/AT1R pathway and the ACE2/Ang (1-7)/Mas receptor pathway (34, 35). Second, it was reported that antibody-dependent enhancement (ADE) of SARS-CoV-2 due to prior exposure to other coronaviruses might also be involved in COVID-19 (36). ADE can elicit sustained inflammation, lymphopenia, and/or cytokine storm, which is a possible explanation for the geographic limitation of severe cases. Third, combined infections may lead to a more severe systemic inflammatory response. Indeed, in our study, some patients had infections in other organs (e.g. urinary tract and blood) caused by other pathogens (e.g., influenza virus and fungi). Fourth, shock, hypoxemia and coagulation pathway abnormalities in critical patients could aggravate the systemic inflammatory response, which lead to a vicious cycle that is life-threatening (37, 38).

In our study, PIRRT was associated with prolonged survival in COVID-19 patients with invasive mechanical ventilation. The primary goal of RRT is to compensate for the loss of renal function and associated sequelae, including uremic toxicity, electrolyte disturbances, metabolic acidosis, and volume overload (22, 39). In addition, RRT can also remove cytokines from the bloodstream. Emerging evidence has shown that RRT was associated with significantly lower mortality in patients with severe sepsis (17, 18, 40). Besides, in patients with acute respiratory distress syndrome, RRT could remove inflammatory mediators, modulate immune function, and regulate oxygenation, thus improving patient prognosis (41-43). PIRRT is a widely used blood purification therapy, that achieves a high solute clearance rate through diffusion and convection (44). PIRRT has shown to encompass the benefits of both continuous RRT in terms of hemodynamic stability and intermittent hemodialysis in terms of cost-efficiency in the intensive care unit (22, 45).

However, it is controversial whether PIRRT is beneficial in viral pneumonia. RRT was reported to have a positive effect on the treatment of adenovirus pneumonia (46). Other studies revealed that PIRRT was a risk factor for mortality in patients with MERS-CoV (15, 47). Yang et al also found that the proportion of nonsurvivors subjected to RRT was higher among patients with COVID-19 (48). In our study, PIRRT was associated with a reduced risk of mortality in COVID-19 patients requiring invasive mechanical ventilation after adjusting for confounding factors. COVID-19 is a novel infectious disease caused by a novel coronavirus, and the underlying pathophysiological process associated with organ involvement is still unclear. In addition, the population studied in our cohort was different from Yang et al., who focused on all critically ill patients, while we focused on patients who received invasive mechanical ventilation. Further research is needed to improve patient treatment and prognosis.

There were some limitations to our study. First, it is retrospective in design, and a prospective double-blind randomized controlled study is warranted in the future. Second, the sample size of this study was not large enough. Third, as it is just a single-center study, multicenter studies are needed for further confirmation.

In summary, we demonstrated that PIRRT could improve COVID-19 patient survival and might be an independent protective factor for COVID-19 patient survival, and might be an independent protective factor for COVID-19 patients with invasive mechanical ventilation. Further prospective multicenter studies with larger sample sizes are required.

## Data Availability

All data that support the conclusions of this manuscript are included within the article.

## Statements

## Acknowledgements

The authors greatly appreciate all the hospital staff for their efforts in recruiting and treating patients and thank all patients involved in this study.

## Statement of Ethics

The study protocol and waiver of written informed consent were approved by the Medical Ethics Committee of Tongji Hospital Affiliated to Tongji Medical College, Huazhong University of Science and Technology (No. TJ-C20200333).

## Author Contributions

F.H., G.X., S.G. and Y.Y. conceived and designed the study. Y.N., J.L, Q.L., S.G., and F.H. were in charge of management of patients. Y.Y., J.S., X.X., Y.W., AC., and F.H. screened, reviewed and recorded the data. Y.Y., J.S. and S.G. performed statistical analyses. Y.Y., J.S. and S.G. drafted the manuscript. All authors provided critical revisions to the manuscript text. All authors read the manuscript and approved the final version.

